# Creating Equity? A Process Evaluation of *Jamkesus Disabilitas*, A Disability-Focused Health Financing Scheme in Yogyakarta, Indonesia

**DOI:** 10.64898/2026.06.29.26356885

**Authors:** Luthfi Azizatunnisa’, Hannah Kuper, Ari Probandari, Lena Morgon Banks

## Abstract

**Background:** This study aims to explore the initiation and implementation of *Jamkesus Disabilitas*, a health financing scheme for people with disabilities in Yogyakarta Province, using the UK Medical Research Council (MRC) Process Evaluation for Complex Intervention.

**Methods:** We interviewed 19 people with disabilities with different types of impairment as beneficiaries, 3 people from Organisations for People with Disabilities (OPDs), 4 government officials, and 4 health providers, either in person, online, or by phone. Interviews were conducted by LA, and took place in Yogyakarta Province in July-September 2024. Data were analysed using a thematic analysis approach using NVivo 15 software.

**Findings:** *Jamkesus Disabilitas* has improved access to assistive technology (AT) and demonstrated inclusive care through its “one-stop service”. It also highlights the importance of consistent leadership in disability-inclusive health systems. However, challenges persist, including uneven AT quality, low coverage, limited availability, and inadequate data for evaluation and planning persisted. Moreover, the absence of inclusive features in the regular service means the scheme has not fully closed the equity gap in healthcare access for people with disabilities.

**Conclusion:** *Jamkesus Disabilitas* has expanded access to AT overlooked by the national health insurance (JKN). However, implementation should prioritise AT quality standards, financial and operational sustainability, and stronger data systems. Broader systemic reforms are also needed to embed disability inclusive practices in regular healthcare service delivery.

## Background

Approximately 16% of world population are people with disabilities [1]. On average, they have higher healthcare needs than those without disabilities, including for general and impairment-related services, as well as assistive technology (AT) [1]. These needs often translate into higher direct healthcare costs, alongside indirect expenses such as transportation, and lost time for both patients and their caregivers [1]. Affordability, therefore, is a substantial barrier for people with disabilities in accessing care, especially since they are more likely to belong to lower socioeconomic groups, have lower capacity to pay, and have lower insurance coverage [2–4]. For example, they are more likely to be uninsured, and many forms of social health insurance offer limited AT coverage or do not cover all healthcare costs incurred [5].

Indonesia hosts *Jaminan Kesehatan Nasional* (JKN – national health insurance), the world’s largest single-payer health insurance scheme with the coverage of approximately 98% of total population in 2024 [6]. Although the coverage amongst people with and without disabilities was similar (65% vs 61%), around 35% of people with disabilities remain uninsured and were less likely to have other insurance coverage than those without disabilities [7]. Discrepancy between these two reported coverages is discussed elsewhere [7]. JKN covers preventive, curative, rehabilitative care, and seven types of AT [8]. While JKN has succeeded in increasing healthcare use for general population, people with disabilities and other vulnerable population [7, 9, 10], its financial protection benefits are uneven. For example, financial protection is pronounced for general population [9]. whereas people with disabilities continue to face substantial out-of-pocket (OOP) spending and experience catastrophic health expenditure (CHE) despite JKN coverage [7]. Furthermore, JKN’s coverage for AT is limited in scope and depth. It covers only seven types of AT and insufficient financial protection, leaving many people with disabilities paying OOP, resorting to substandard AT, or forgoing essential AT, resulting in unmet needs [11]. A previous study showed that up to 60% people in need of AT in Indonesia do not receive it [11].

*Jamkesus Disabilitas* is a provincial-level disability-focused health financing scheme in the Special Region of Yogyakarta. It was established in 2013 (i.e., before the launch of JKN), and it initially served as a comprehensive provincial health insurance scheme, covering promotive, curative, and rehabilitative services. Following the establishment of JKN as a single-payer system, its role has shifted into a complementary, *add-on* scheme designed to address gaps in national coverage, particularly through the provision of AT and other disability-related services such as homecare for pressure sores. To be eligible for *Jamkesus Disabilitas*, individuals must be residents with disabilities and either lack health insurance or have disability-related health needs that are not covered under their existing health insurance. This initiative demonstrates how subnational governments can advance health equity for people with disabilities and offers valuable lessons for other settings. Therefore, this study aims to explore the initiation and implementation of *Jamkesus Disabilitas* using the United Kingdom Medical Research Council (UK MRC) Process Evaluation for Complex Intervention Framework [12].

### Yogyakarta and Jamkesus Disabilitas

The Special Region of Yogyakarta is an autonomous province on the Island of Java, Indonesia, uniquely ruled by the Yogyakarta Sultanate, with the Sultan serving as governor. The special region status grants autonomy to implement *Jamkesus Disabilitas* and have its managing body without national approval. The province is relatively small with only four districts and one city. The population in 2025 is approximately 3.7 million [13], with around 10% population living in poverty [14]. There is an estimated 26,371 people with disabilities (0.6% of the population) in 2024, although this is likely to be a severe underestimate [15].

Yogyakarta hosts many Organisations of Persons with Disabilities (OPDs) working in areas such as advocacy, employment, education, and health. In 2009, these OPDs formed the Indonesian Disability Network (IDN) following the 2006 earthquake, which left many survivors with new disabilities [16, 17]. The IDN successfully advocated for a health insurance scheme to address the needs of people with disabilities who lacked coverage. Since then, OPDs in Yogyakarta have remained central to promoting empowerment and advocating disability rights [16].

In 2011, the provincial government of Yogyakarta introduced the Universal Health Insurance (*Jaminan Kesehatan Semesta* – *Jamkesta*) to cover Yogyakarta residents (See the timeline in Figure 1). *Jamkesus Disabilitas* was established in 2013 as a health insurance scheme to provide continuous and affordable healthcare for persons with disabilities. It initially used a membership system (i.e., contributory and subsidised), in which beneficiaries received a card to access services. In 2014, its benefits were expanded to include AT. Following the introduction of JKN in the same year, mandated as the country’s sole national health insurance, *Jamkesus Disabilitas* adapted its role. By 2017, it had gradually transitioned into an “add-on” scheme, which remains in place to the present day. The membership mechanism, then, was discontinued. There is no longer contribution as the implementation is fully funded by the government. Beneficiaries are no longer enrolled as members; instead, they must register and undergo verification for each service use. Verification confirms residence, self-reported disability status (without requiring certification), and that the requested healthcare or AT is not covered by JKN. Over time, the scheme’s services have increasingly focused on AT provision. A comparison of AT coverage between *Jamkesus Disabilitas* and JKN is shown in Supplementary 1.

*Jamkesus Disabilitas* covers preventive, curative, and rehabilitative as shown in Box 1 [18]. The services operate through a tiered referral system (regular system), requiring multiple procedures and institution visits for registration, verification, and care, creating high indirect costs (Figure 2). Initially, this was the only option, but its complexity led to low utilisation. In response, *Jamkesus Disabilitas* Administering Body (*Balai Penyelenggara Jaminan Kesehatan Sosial* – *Bapel Jamkesos*) introduced “one stop service”, integrating administrative and service delivery in one location (Figure 2). The “one-stop service” provides screenings (i.e., for NCDs, pressure sores – common among wheelchair users, and TORCH – often cause congenital impairment), consultation with general practitioners, physiatrists, and AT service, with referrals as needed. The majority of beneficiaries access AT through “one-stop service”, except for hearing aid, which remains in the regular system due to the need for specialised testing in a quiet room. The “one-stop service” is an example of how inclusive healthcare service may look like; it is held in an accessible venue (e.g., equipped with accessible toilet, ramps, wheelchairs provided), transportation is provided, escorts assistance, and healthcare workers are experienced to provide services for people with disabilities.

Initially, the “one-stop service” was organised by *Bapel Jamkesos* and funded by the Provincial Government. Since 2018, district governments have taken over organising and funding activities such as covering venue, transportation, escorts and assistance, volunteers, and community mobilisation, while healthcare and AT services remain funded by *Bapel Jamkesos*. Each district holds at least one “one-stop service” annually, plus one by Bapel Jamkesos, totalling at least six events per year. In 2023, *Jamkesus Disabilitas* was integrated into the Universal Health Insurance (Jamkesta - Governor Regulation No. 7/2023), and expanding eligibility from “people with disabilities and poor” to only “people with disabilities”.

**Box 1.**
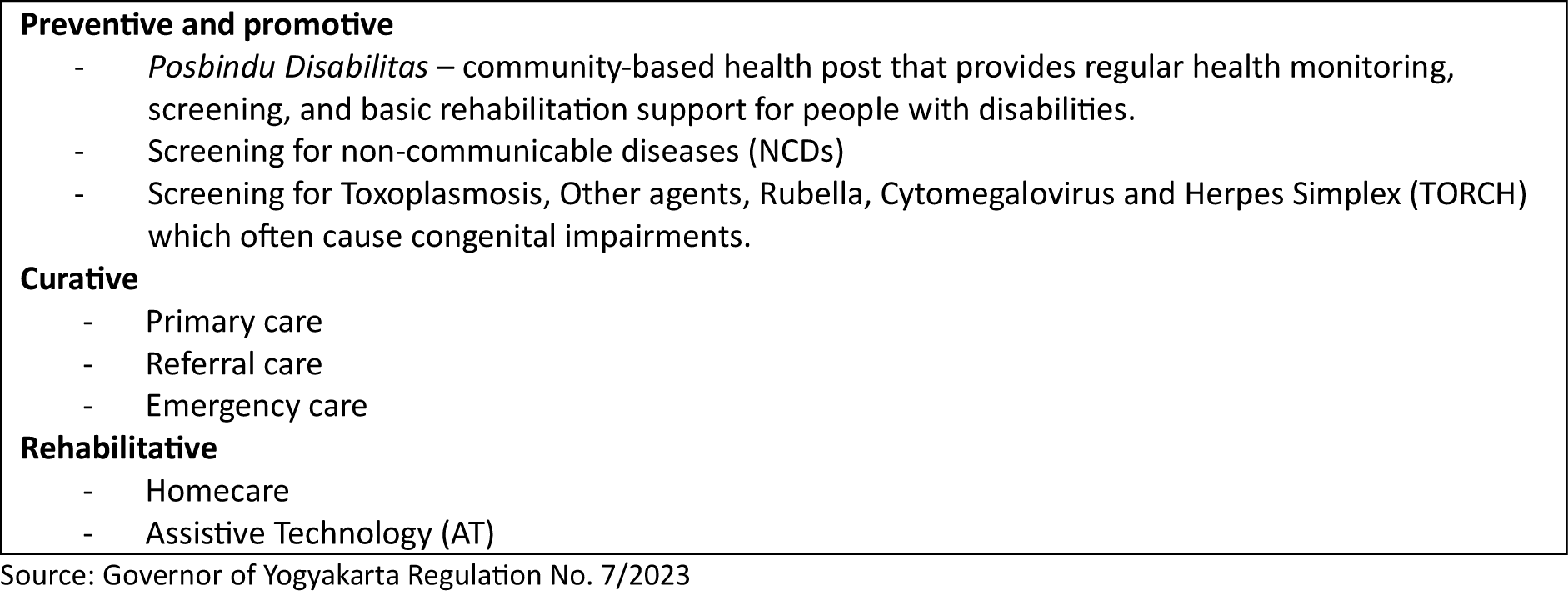
Services Covered by *Jamkesus Disabilitas* (Disability-Focused Health Financing) in Yogyakarta Province, Indonesia.

## Methods

### Study design

This study is a process evaluation study using qualitative methods to explore the implementation of *Jamkesus Disabilitas* in Yogyakarta Province, Indonesia. Data collection was conducted in July - September 2024.

### Participants and recruitment

This study involved two groups of participants: people with disabilities who were beneficiaries of *Jamkesus Disabilitas*, and key informants involved in the programme’s implementation.

People with disabilities were recruited to explore their experiences with accessing the scheme. Recruitment considered diversity in terms of impairment type, sex, and area of residence (urban or rural) to capture a wide range of perspectives. A snowball sampling strategy was applied. The initial list of potential participants was obtained through the main author (LA) and the local OPDs network. These initial participants were then asked to suggest the next potential participants.

Key informants were recruited from three groups to gain insights of the implementation of *Jamkesus Disabilitas*, i.e., government officials, service providers (i.e., physicians and AT providers), and OPDs. Snowball sampling strategy was also employed. Initial contacts were identified through the Provincial Government and OPDs networks, who then referred other relevant stakeholders.

### Data collection procedures

Data were collected through in-depth interviews with both beneficiaries and key informants. Interviews were conducted in different formats depending on the participants’ preferences and accessibility needs: in-person, by phone, or online via video conferencing (particularly when participants required a sign language interpreter). Informed consent was obtained prior to each interview, which was written for in-person, and verbal (audio-recorded) for telephone and online interviews. Participants were informed about the purpose of the study, voluntary participation, confidentiality, potential risk, and their right to withdraw at any time. All interviews were audio-recorded with participants’ permission, and field notes were taken during and after the sessions to capture contextual details and initial impressions.

Interviews were conducted in the Indonesian Language and guided by semi-structured interview guides. These guides were developed using the UK MRC Process Evaluation Framework (Supplementary 2) [12], and were adapted for different categories of participants (Supplementary 3).

### Data analysis

Data were analysed thematically, guided by the UK MRC process evaluation framework for complex interventions [12]. This framework provided a set of predetermined themes, including context, implementation, mechanisms of impact, and impact. In addition, inductive analysis was applied to capture emerging themes, such as sustainability, that extended beyond the original framework.

The analysis followed several steps. First, interview recordings were transcribed verbatim by an experienced transcriber and checked by LA. She then read through all transcripts to become familiarised with the data. Next, coding was conducted using NVivo 15. Codes were then grouped into subcategories and categories, which were further organised into overarching themes. The themes were discussed with other team members. The themes were mapped in Figure 3.

Several strategies were used to enhance the reliability of the study findings. Triangulation was achieved by collecting data from multiple groups i.e., beneficiaries, government representatives, service providers, and OPDs, allowing comparison and validation across perspectives. Member checking was conducted with government representatives and OPDs, where interim results and emerging themes were shared to confirm accuracy and relevance. Peer debriefing involved LA summarising each interview and discussing initial themes with the research team in regular meetings, which supported iterative interpretation, and reduced individual bias.

### Research team and reflexivity

All interviews and preliminary analysis were conducted by LA, a PhD researcher in epidemiology and population health with substantial qualitative research experience, and an Indonesian woman with disabilities, whose lived experience informed the research process and engagement with participants. Interpretation and analysis were further discussed with HK, AP, and LMB to enhance reflexivity and minimise individual bias in interpretation. They are all women. HK and LMB are based in the UK and have extensive experience in global health and research on disability, while AP is based in Indonesia and has extensive experience in health system research in Indonesia.

### Ethical approval

Ethical approval was obtained from the Research Ethical Committee of the London School of Hygiene & Tropical Medicine (LSHTM) (Ref no: 30136) and the Faculty of Medicine, Public Health and Nursing, Universitas Gadjah Mada, Indonesia (Ref no: KE/FK/0410/EC/2024).

## Results

### Characteristics of Participants

We approached 22 potential participants with disabilities, and successfully interviewed 19, including people with physical, vision, hearing, psychosocial, and multiple impairments. Three potential participants were excluded due to communication difficulties or geographical inaccessibility. Additionally, we interviewed 11 key informants from government, OPDs, and healthcare providers (Table 1). Among participants with disabilities, the majority were women, adults, had physical impairment, and were either unemployed or engaged in the informal sector.

**Table 1.** Characteristics of Participants.

| Characteristics of participants | N (%) |
| --- | --- |
| Type of participants |  |
| People with disabilities (beneficiaries) | 19 (63%) |
| OPDs | 3 (10%) |
| Government | 4 (13%) |
| Health providers | 4 (13%) |
| **Characteristics of people with disabilities (N:19)** | **N (%)** |
| Sex |  |
| Female | 12 (63%) |
| Male | 7 (37%) |
| Age |  |
| <18 | 4 (21%) |
| 18-60 | 14 (74%) |
| >60 | 1 (5%) |
| Impairment types |  |
| Physical | 8 (42%) |
| Vision | 1 (5%) |
| Hearing | 5 (26%) |
| Psychosocial | 2 (11%) |
| Multiple | 3 (16%) |
| Residence |  |
| Urban | 9 (47%) |
| Rural | 10 (53%) |
| Work status |  |
| Student | 5 (26%) |
| Self-employed/informal | 5 (26%) |
| Work for others | 3 (16%) |
| No work | 6 (32%) |

### Context – Factors influence the initiation and implementation of Jamkesus Disabilitas

The establishment of *Jamkesus Disabilitas* was primarily driven by strong government commitment, shaped by heightened awareness of disability issues and concern for poor and vulnerable populations. The 2006 earthquake, which increased the number of people with disabilities and attracted international aid, raised government responsiveness to the needs of people with disabilities. At the same time, sustained advocacy from OPD alliances kept disability on the policy agenda. The commitment was then translated into regulations and budget allocation.

OPDs advocacy strategies included involving government officials in OPDs programmes to raise awareness and build support, collaborating with academics to generate evidence, coordinating advocacy across OPDs, leveraging media to generate public pressure, and framing disability as a political and electoral issue to attract attention from both local government and politicians. Beyond advocacy, the meaningful involvement of OPDs in programme planning, implementation, and evaluation was also pivotal in shaping *Jamkesus Disabilitas*. They also played a crucial role in generating data to target beneficiaries, which served as the foundation for the programme’s initiation.

Yogyakarta’s special regional status further facilitated this process. Its autonomy enabled the establishment of a dedicated provincial body (*Bapel Jamkesos*) under the Provincial Health Office to implement and manage *Jamkesus Disabilitas*. Over time, the programme gained national and international recognition, positioning Yogyakarta as a pioneer in disability-inclusive health policy. The recognition further reinforced the provincial government’s commitment to retaining *Jamkesus Disabilitas* after the establishment of JKN.

> “*The key factor is the government’s commitment. We are fortunate to have a governor who prioritises the needs of the vulnerable population. Since Jamkesus Disabilitas is under the Sultan—who has never been replaced—the policy has remained a consistent priority to protect vulnerable populations*.*” (Man, without impairment, OPD)*

### Implementation: Reach and Coverage, Dose, Fidelity, and Adaptation

#### Reach and Coverage

The utilisation of *Jamkesus Disabilitas* services increased substantially after the launch of the “one-stop service” in 2015, as most beneficiaries accessed services through this mechanism. However, utilisation declined sharply in 2020 due to COVID-19 restrictions on mass gatherings, and post-pandemic use has remained lower than pre-pandemic (Figure 4).

*Bapel Jamkesos* adopted a range of outreach strategies to extend coverage and improve accessibility. These strategies included distributing individual invitations to prospective beneficiaries, collaborating with social workers and OPDs to identify potential beneficiaries, engaging with volunteers and village governments for community mobilisation, and providing transport and escort services. Outreach was further strengthened by involving non-health actors such as *Babhinkamtibmas* (community police officers) and community organisations to reach people with disabilities who might not otherwise be identified.

Despite these efforts, challenges persist. Reliable disability-related data are scarce, hindering accurate identification of beneficiaries and limiting the ability to assess programme coverage, as one of the government officials explained:

> *“The main challenge for us is data. We don’t know exactly how many people with disabilities are in Yogyakarta—only that there are around 29 to 30 thousand. We don’t have names or addresses, we can’t assess what types of disabilities exist and how many we should cover” (Woman, a government official)*

Coverage is likely, however, to be inadequate. Between 2018 and July 2025, *Jamkesus Disabilitas* dispersed 1,677 adaptive wheelchairs, 174 hearing aids, 348 spectacles, and 1,298 orthoses and prostheses (Figure 5). Applying national prevalence estimates to Yogyakarta’s population of 3.7 million in 2025, AT distributed by *Jamkesus Disabilitas* remains far below the estimated need and access (Table 2) [11].

**Table 2.**
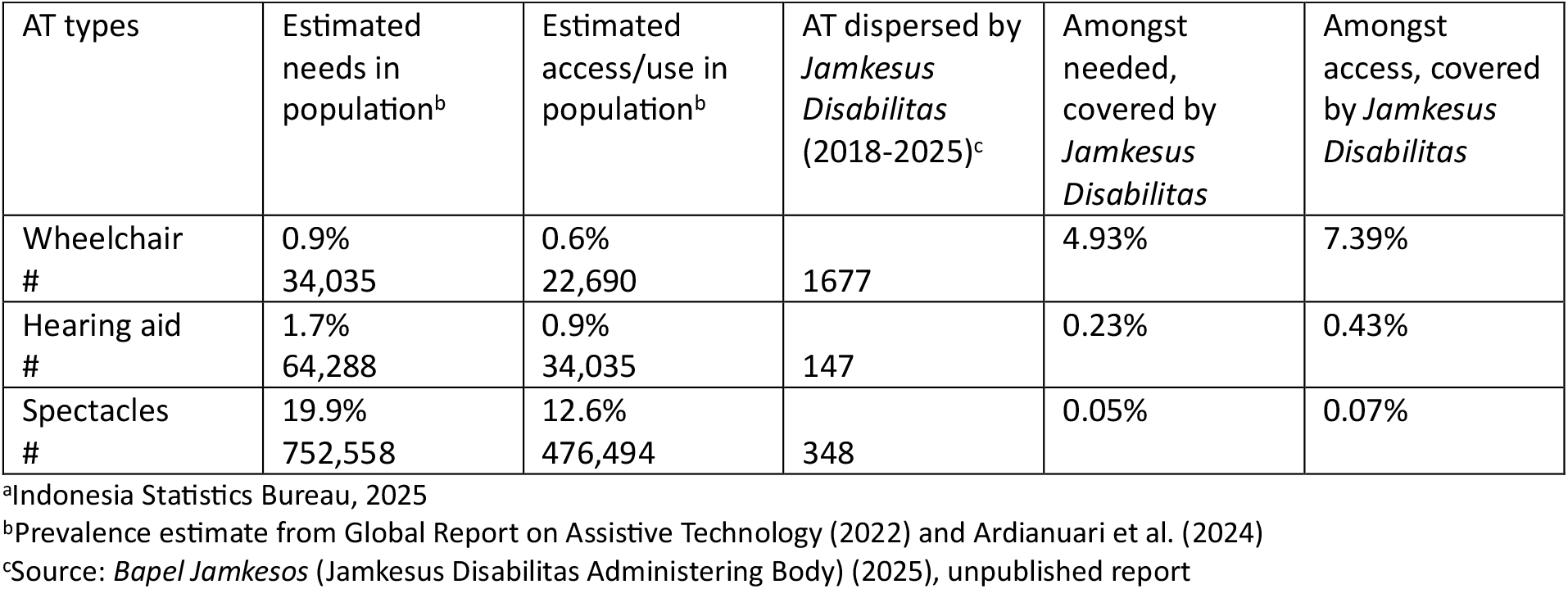
Estimated Coverage of Assistive Technologies (AT) Distributed by *Jamkesus Disabilitas* (Population in Yogyakarta in 2025: 3,781,700^a^)

Access to information on the scheme was variable. Participants predominantly identified OPDs, community networks, social workers, and social media as the primary sources of information about *Jamkesus Disabilitas*. Nonetheless, notable information gaps persisted among beneficiaries. Lack of information was particularly common amongst individuals with psychosocial impairments, as *Jamkesus Disabilitas* was widely perceived primarily as a source for AT with limited services for individuals with psychosocial conditions.

#### Dose

The “one-stop service” is scheduled at least six times a year across Yogyakarta. Despite this arrangement, participants reported irregular scheduling as a barrier to participation. Another concern is that “one-stop service” cannot accommodate urgent AT services. Beneficiaries often have to wait until the next event, rely on regular tiered referral services, pay OOP or forgo the needed service.

> *“Often, when I need wheelchair repair, it just happens not to coincide with the “one-stop service” schedule*.*” (Woman, physical impairment, beneficiary)*

To mitigate geographic and scheduling barriers, beneficiaries are allowed to attend services in other districts, offering greater flexibility and extending access beyond their immediate locality.

Nevertheless, this concern persisted and was echoed by other beneficiaries, OPDs, and officials, who noted that *Jamkesus Disabilitas* has become more focused on the periodic “one-stop service” event, rather than strengthening regular services to ensure timely, routine, and simple procedures. Although regular mechanisms remain available, many people avoid them due to the complex procedures and referral processes that require multiple visits and cause high indirect costs.

> *“Now the focus is more on the “one stop service”. Actually, the original concept was that assistive devices could actually be accessed on a daily basis, without having to wait for a three-monthly event*.*” (Man, physical impairment, OPD)*

#### Fidelity

The “one-stop service” was designed to enhance access to healthcare and AT services. Its implementation is largely consistent with the initial plan, incorporating accessibility features such as ramps and accessible toilets, along with support measures including transportation services (e.g., ambulances for pick-up) and escort teams to assist beneficiaries. Several components need further refinement to ensure equitable and sustained access, for example registration.

The “one-stop service” provides online registration only, which was perceived as a barrier, particularly by those with vision impairment, limited digital literacy, or no access to a device. Although social workers and OPDs offer assistance, these measures have not fully addressed the barriers, underscoring the need for more inclusive registration options.

> *“The requirements, like taking and uploading photos via Google Form, were difficult for blind people. Many don’t have a smartphone, and some just give up*.*” (Man, vision impairment, OPD)*

Accessibility challenges also affect the fitting and collection of prostheses, which require travel to AT providers. Distance and transport barriers leave many devices unclaimed, preventing providers from securing reimbursement. In response, collective distribution via “one-stop services” was introduced. However, a new challenge emerged: prostheses fitting requires bulky equipment that is difficult to transport, and in some cases, not all necessary tools can be brought to each location, resulting in suboptimal fitting. Additionally, the equipment requires a high electricity capacity, which is often a constraint in the field.

Although regulations specify a wide range of AT with higher financial coverage than JKN (Supplementary file 1), in practice, the provision remains limited, primarily to adaptive wheelchairs, certain types of orthoses, and prostheses (e.g., upper- and lower-limb prostheses). Low-vision spectacles, for example, were previously available, but difficulties in conducting proper assessments and the scarcity of qualified service providers restricted access. For many other types of AT, especially the wide variety of orthoses and prostheses, it remains unclear whether the low utilisation reflects limited service availability, prescribing practices, the absence of eligible users, or lack of awareness among potential beneficiaries about the range of AT that can be accessed through *Jamkesus Disabilitas*.

Additionally, the quality of devices varies across providers. Both beneficiaries and the government reported inconsistencies in durability, comfort, and suitability across different types of AT. These variations raise concerns about whether *Jamkesus Disabilitas* was delivering on its intended goal of reliable and quality AT. The absence of a clear quality standard likely contributes to these disparities, which in turn may lead to dissatisfaction, abandonment, or underuse of AT, potentially limiting the intended benefit of AT for users.

> *“Our challenge in AT provision is that some aspects are still not optimal, and the quality still needs improvement” (Woman, a government official)*

#### Programme adaptation

Three major adaptations of *Jamkesus Disabilitas* were identified since its launch. First, the innovation of “one-stop service” was a response to the low use of regular services due to complex administrative procedures and barriers related to distance and transportation. Its introduction faced resistance within the Provincial Health office, as it challenged rules requiring services to be delivered at health facilities and strict health insurance regulations on verification procedures. The initiative’s budget was initially not approved. Consequently, implementers and OPDs advocated to the provincial parliament and governor, engaged local politicians, involved the press, and mobilised crowdfunding and private-sector donations. With this public pressure and the argument that the “one-stop service” would overcome the bottleneck of utilisation, it gained stronger government backing, eventually leading to formal regulation and dedicated budget allocation.

Second, the service was expanded to cover homecare for pressure ulcers due to the high frequency among spinal cord injury cases, and TORCH screening, following strong advocacy from the cerebral palsy community. However, the TORCH screening lacked accompanying follow-up education, resulting in knowledge and awareness gaps.

> *“What’s missing is continuity — after TORCH screening, parents should receive follow-up education so they can learn and manage independently*.*” (Woman, caregiver of an individual with multiple impairments [19], beneficiary & OPD)*

Third, *Jamkesus Disabilitas* evolved from a comprehensive health insurance into an add-on financing scheme. After the rollout of JKN, local schemes were expected to be phased out. Nevertheless, the Yogyakarta provincial government retained *Jamkesus Disabilitas* to address disability-specific needs and fill the gaps in JKN coverage.

> *“The Sultan remained consistent in protecting the poor and vulnerable population. Despite national pressure to merge local schemes into JKN, he refused to dissolve Jamkesta and ensured Jamkesus Disabilitas continued, even expanding coverage to over 50 types of assistive devices*.*” (Man, individual with physical impairment, OPD)*

### Acceptability and perceived impact

Beneficiaries reported that *Jamkesus Disabilitas* provided a degree of financial protection by covering the costs of AT. However, additional expenses for the repair and maintenance remained a financial burden for many. Beneficiaries perceived that the programme also facilitated ongoing health monitoring with regular health screening services serving as important mechanisms for early detection and continuity of care.

Improvements in quality of life (QoL) were frequently mentioned by interviewees. The provision of hearing aids, for instance, enhanced communication and social interaction, while access to mobility devices increased independence, participation in daily activities, and self-confidence. These outcomes reflected not only the functional benefit of AT but also its social and psychological significance.

Experiences with AT and related services by users were mixed. Many beneficiaries expressed satisfaction, highlighting the usefulness of devices and appreciation for the support received. Others, however, reported frustration and disappointment when devices were ill-fitting or inadequate for their needs.

> *“The hearing aids from provider [A] are too weak, don’t fit his needs, and don’t have an earmold. I wish he could get ones like provider [B]’s — I saw my son’s friend using them, and they’re much better with proper earmolds*.*” (Woman, caregiver of a child with hearing impairment [6], beneficiary)*

Despite these concerns, opportunities to voice dissatisfaction were limited. Evaluation led by *Bapel Jamkesos* was perceived as hierarchical, creating a power imbalance that discouraged open feedback. Many beneficiaries still experience confusion, fear, or reluctance to file complaints, indicating gaps in awareness, accessibility, and trust in the existing mechanisms.

> *“Bapel Jamkesos did the evaluations by going directly to the users and asking if they’re satisfied with the service. There’s a power imbalance. People often feel awkward responding and don’t speak up. It’s only in discussions without Bapel Jamkesos that they admit the ATs aren’t good in quality*.*” (Man, physical impairment, OPD)*

At the systems level, *Jamkesus Disabilitas* had broader impacts. The programme gained recognition at both national and international levels, positioning Yogyakarta as a benchmark for disability-inclusive health initiatives. This recognition spurred interest from other provinces and the national government in adopting similar models. Additionally, *Jamkesus Disabilitas* strengthened the delivery of inclusive healthcare services and raised awareness among government officials, healthcare providers, and the wider community about the specific health needs of people with disabilities.

### Sustainability

Sustainability emerged as a cross-cutting theme and was subsequently incorporated into the framework as an additional component (indicated by a dashed line in Figure 3).

*Jamkesus Disabilitas* was integrated into *Jamkesta* (Universal Health Insurance) to promote long-term viability, aiming to streamline institutional structures, mainstream disability services, and ensure the sustainability of these efforts. The eligibility criteria were also broadened from poor individuals with disabilities to all residents with disabilities, enhancing inclusivity by recognising disability itself as a basis for protection.

From a financing perspective, *Jamkesus Disabilities* has transitioned from relying solely on provincial funding to a co-funding mechanism with district governments. However, sustainability challenges persist in the provision of AT. While prostheses are produced locally by contracted providers, adaptive wheelchairs remain dependent on foreign aid or imports, incurring high tariffs that providers must absorb even when devices are donated. This reliance on external aid poses long-term risks, as any reduction in support could undermine continuity. In 2025, wheelchair providers began exploring collaborations with local manufacturers to reduce dependence on imports. Additionally, *Bapel Jamkesos* requires a clear plan to enhance affordability, reduce reliance on external aid, and ensure the continuity of AT provision, including advocating for domestic AT manufacturing and national-level policy reforms to regulate or eliminate tariffs.

## Discussion

*Jamkesus Disabilitas* is a disability-focused health financing scheme in Yogyakarta Province that seeks to address the health needs of people with disabilities, particularly on access to AT. Its key features offer lessons including strong and consistent government commitment through policies and regulations, cross-sectoral collaboration, meaningful participation of OPDs and their sustained advocacy. It also showcased disability-inclusive healthcare delivery through its “one stop service” and adaptation of the programme to the needs of people with disabilities, including flexible eligibility criteria, and alignment with the universal and mainstream health insurance system. However, important challenges remain. Importantly, gaps remain in terms of AT quality, availability, and coverage. There is also a need to strengthen data on disability and feedback mechanism for evaluation. A clear sustainability plan, especially for AT provision and financing, is also critical to ensure the programme’s long-term viability.

The provision of AT and related services can enhance equity in AT access. This provision is particularly important in LMICs where mainstream health insurance schemes often either exclude AT or provide only limited coverage [5, 19, 20]. As a result, people with disabilities frequently rely on OOP payments or donations to obtain AT [11, 21], leaving many with unmet needs or high costs [22]. Improved access to AT can help address these unmet needs and promote participation (e.g., education and employment), independence, quality of life and safety as recognised in the Convention on the Rights of Persons with Disabilities [23, 24].

*Jamkesus Disabilitas* applies flexible eligibility criteria, allowing people with self-reported disability, with no certification required, to access its services. This approach can help eliminate the cumbersome, lengthy, and costly medical-based assessment procedures, which often restrict access to social protection in many countries, paradoxically excluding the population such programmes are intended to support [20, 25–27]. It also aligns with recommendations from the UN Committee on the Rights of Persons with Disabilities, which advocates shifting disability determination mechanisms from a medical model to a human rights model, applying disability assessment which is accessible, affordable, and minimally burdensome [28].

*Jamkesus Disabilitas* does not impose a poverty criterion for eligibility. This approach is more disability-inclusive, as it bases access on disability-related needs rather than income status. Many social protection programmes in LMICs are primarily embedded within poverty-reduction frameworks, which exclude many people with disabilities who do not meet strict poverty thresholds despite facing substantial disability-related costs [20, 25–27]. Focusing on needs is usually better because relying mainly on means or proxy-means testing often results in greater inequality [29].

Is *Jamkesus Disabilitas* creating equity? The scheme has improved access to AT, yet coverage, availability and quality are still uneven. The “one-stop service” demonstrates what inclusive care can look like with an accessible venue, provision of transport and human assistance, and providers trained in disability-inclusive practice. However, the one-stop services are held irregularly and this level of inclusion is not embedded in the regular service delivery mechanism, where complex procedures and high indirect costs continue to limit use. As a result, it has not fully closed the equity gap in healthcare. Achieving true equity will require systemic reform that extends inclusive practices beyond *Jamkesus Disabilitas* and into mainstream health services, including JKN [30].

*Jamkesus Disabilitas*, however, highlights the importance of consistent leadership and political commitment to advancing disability-inclusive health. In Yogyakarta, the 2006 earthquake created political momentum to address the needs of people with disabilities, catalysing the development of *Jamkesus Disabilitas*. Strong advocacy from local OPDs further shaped its design and sustained attention to disability issues. This experience demonstrates how political will can elevate neglected issues, mobilise cross-sectoral actors, and secure the resources needed to implement and intain inclusive health initiatives [31, 32].

Persistent debate over whether disability-related needs are best addressed through targeted or mainstream health financing/insurance occurs. Targeted insurance schemes can specifically address the needs of people with disabilities, but face sustainability challenges, particularly when serving populations with higher health needs and limited financial capacity [33]. In contrast, universal schemes can promote rights, impartiality, and greater potential for scaling up [29, 34]. Yet, universality alone often fails to meet the diverse needs of people with disabilities [29]. A twin-tracked approach, integrating universal inclusion with targeted, disability-specific support, could serve as an intermediary step to incorporate impairment-related health service and AT into the mainstream health insurance scheme.[35] The integration of *Jamkesus Disabilitas* into *Jamkesta* is an example of how targeted schemes can evolve toward mainstream inclusion while maintaining disability responsiveness.

The study findings have implications for policy and practice. First, addressing the issues of AT quality and availability requires advocacy at the national level to strengthen supply chains, and enhance cross-sector coordination. Key strategies include developing national action plans and policies, shaping and enabling markets through incentives for local manufacturing to produce affordable and contextually appropriate devices, and strengthen the workforce capacity.[34] Second, strengthening data systems is essential for the evaluation and continuous improvement of *Jamkesus Disabilitas* and similar interventions. Third, adopting *Jamkesus Disabilitas* elsewhere in Indonesia requires adaptation rather than direct replication. Understanding context (e.g., organisational capacity, resources, social support, leadership) is vital, as interventions effective in one setting may not work in another [36–39]. Finally, inclusive features demonstrated by *Jamkesus Disabilitas* through the “one-stop service” should be embedded into mainstream healthcare services to improve equity in access.

This study’s strength is that this study incorporates perspectives from a wide range of stakeholders, including government actors, healthcare and assistive technology providers, OPDs, and beneficiaries with diverse impairments. Additionally, it explores *Jamkesus Disabilitas* comprehensively from initiation to implementation. It does, though, have several limitations. First, reliance on qualitative methods may introduce researcher bias, although reflexivity, triangulation, debriefing, and member checking were used to enhance rigour. Second, recruitment through snowball sampling via OPDs and LA’s networks may have excluded individuals not connected to these groups, particularly those hardest to reach. People involved in OPDs may differ from those not affiliated with such organisations, as they tend to be more informed, which may shape their access to services and engagement with health and social programmes. Third, we were unable to include individuals with severe communication barriers, even when reasonable adjustments such as sign language interpreter or proxy interview were provided. As a result, people with these characteristics may be underrepresented in this study. Finally, data collection was constrained by accessibility barriers, with some potential participants unable to take part due to environmental inaccessibility or limited use of communication devices, highlighting the very challenges people with disabilities face in accessing healthcare.

## Conclusion

*Jamkesus Disabilitas* has enhanced access to AT, and is supported by several key facilitators, including government commitment, multisectoral collaboration, continuous advocacy, and meaningful participation of OPDs, and programme adaptation to meet the needs of people with disabilities. However, AT quality across providers should be improved through the establishment of quality standards and enhanced monitoring and evaluation, and the sustainability of AT provision needs to be ensured through long-term planning.

## Acknowledgement

The authors wish to thank all study participants for their valuable contributions, the transcriber for their assistance, and the funding body for its support

## Author contribution

LA conceptualised and designed the study with LMB and HK’s input. LA prepared the protocol, conducted data collection and analysis, interpreted the findings, and drafted the manuscript. HK and LMB contributed by reviewing the protocol, interpreting the results, and revising the manuscript. AP also supported interpretation and manuscript review. All authors reviewed and approved the final version and take responsibility for its content

## Data availability

The authors can provide anonymised interview transcripts upon reasonable request.

## Declarations

### Ethics approval and consent to participate

Ethical approval was obtained from the Research Ethical Committee of the London School of Hygiene & Tropical Medicine (LSHTM) (Ref no: 30136) and the Faculty of Medicine, Public Health and Nursing, Universitas Gadjah Mada, Indonesia (Ref no: KE/FK/0410/EC/2024). Informed consent was obtained prior to each interview: written consent for in-person interviews, and verbal, audio-recorded consent, for telephone and online interviews. Participants were informed about the study’s purpose, the voluntary nature of participation, confidentiality, potential risk, and their right to withdraw at any time before agreeing to participate. All procedures adhered to ethical standards in accordance with the Declaration of Helsinki.

### Consent for publication

Not applicable

## Declaration of generative AI and AI-assisted technologies in the writing process

LA drafted the manuscript and used AI (ChatGPT) to check the grammar, and improve language and readability of the text. All authors then reviewed and edited the content as needed and took full responsibility for the content of the publication.

## Abbreviation

AT: Assistive Technology
Bapel Jamkesos: Balai Penyelenggara Jaminan Kesehatan Sosial – Jamkesus Disabilitas administering body
IDN: Indonesian Disability Network
OOP: Out-of-pocket
OPDs: Organisation of People with Disabilities
QoL: Quality of Life
CHE: Catastrophic Health Expenditure
TORCH: Toxoplasmosis, Other agents, Rubella, Cytomegalovirus, Herpes Simplex Virus
NCDs: Non-Communicable Diseases
JKN: Jaminan Kesehatan Nasional – National Health Insurance Scheme
LMICs: Low- and Middle-Income Countries
UK MRC: United Kingdom Medical Research Council
UN: United Nations

