## Supplementary Files for "Creating Equity? A Process Evaluation of *Jamkesus Disabilitas*, A Disability-Focused Health Financing Scheme in Yogyakarta, Indonesia"

Supplementary File 1. Gaps of Assistive Products Coverage between the National Health Insurance (JKN) and *Jamkesus Disabilitas*

| **No** | **Type/name of AT** | **JKN** | **Jamkesus Disabilitas** |
| --- | --- | --- | --- |
| 1 | Spectacles | - Provided at a minimum interval of once every two years. - Medical indications: at least – Spherical 0.5D, Cylindrical 0.25D. - Provided based on a prescription from an ophthalmologist. - Tariff in IDR: 165,000 – 330,000 | - Single Vision CR-39 with Frame – Range Sph+CYL -10 to Sph+CYL +6.00. Costs (IDR): from 390,000 to 500,000 - CR-39 Bifocal Kryptok with frame   HMC Sph+CYLMaks +2.00 s/d-2.00: max cost of IDR 630,000  HMC RX Sph+CYL Maks +7.50 s/d -10.00: max cost of IDR 550,000  LENTICULER OMEGA HMC Sph+CYL Maks +7.00 s/d+17.00: max cost of IDR 1,250,000   - Low vision spectacles Sph+CYL (SV LENTICULAR HMC) up to -25 and +25. Max cost of IDR 1,250,000 – 1,650,000 |
| 2 | Low vision devices | None | Many types of magnifiers. Max cost range IDR 950,000 – 1,550,000 |
| 3 | White cane | None | Maximum cost of IDR 250,000 |
| 4 | Hearing aid | - Provided at a minimum interval of once every five years, based on medical indications, regardless of whether for one or both ears, and for the same ear. - Provided upon prescription from an ENT specialist. - Maximum cost of IDR 1,100,000. | - 0 – 24 db: 0 (no need hearing aid) - 25 – 90 db: max cost of IDR 4,000,000 - 91 – 120 db: max cost of IDR 7,500,000 - >120 db: hearing loss (no need hearing aid) |
| 5 | Protheses | - **Prosthetic devices for mobility include: a**rtificial leg and artificial arm Max cost of IDR 2,750,000 - Provided at a minimum interval of once every five years, based on medical indications for the same type of prosthetic device. | Covering 36 types of protheses   - Lowest price: IDR 1,440,000 wrist disarticulation prostheses - Highest price: IDR 18,890,000 hip disarticulation prostheses endoskeletal system |
| 6 | Teeth protheses | - Provided at a minimum interval of once every two years, based on medical indications for the same teeth. - Full dental prosthesis: maximum cost of IDR 1,100,000. - Each jaw: maximum cost of IDR 550,000. | None |
| 7 | Orthosis – braces | Spinal corset - Provided at a minimum interval of once every two years, based on medical indications, with a maximum cost of IDR 385,000 | Cover wide range of corsets and braces  Trunk orthoses:   - Lowest price: elastic corset IDR 189,000 - Highest: corset orthoses polyetheline IDR 4,700,000   Upper limb orthoses   - Lowest: hand splint IDR 405,000 - Highest: airplane splint IDR 2,533,800   Leg orthoses   - Lowest: Rigid back slab IDR 322,000 - Highest: Hip knee ankle foot orthoses IDR 5,687,000   Foot orthoses   - Lowest: Donut heel IDR 145,000 - Highest: Standing frame IDR 4,620,000 |
| 8 | Orthosis – collar neck | Provided at a minimum interval of once every two years, based on medical indications, with a maximum cost of IDR 165,000. | - Hard cervical collar: IDR 653,000 - Soft cervical collar: IDR 181,500 |
| 9 | Crutches | Provided at a minimum interval of once every five years, based on medical indications, with a maximum cost of IDR 385,000. | Wide range of walking aids   - Under arm crutches: IDR 256,000 - Canadian crutches: IDR 197,000 - Tripod: UDR 197,000 - Different types of walkers, range IDR 531,000 – 800,000 - Quad cane: IDR 197,000 - Walking stick: IDR 197,000 |
| 10 | Wheelchairs | None | Standard wheelchair IDR 1,485,000  Wide range of adaptive wheelchairs   - Lowest cost: 4,620,000 - Highest: autism/cerebral palsy/hydrocephalus (complex rehabilitation): IDR 31,845,000   Maintenance service: IDR 27,000 – 110,000  Wide range of spare parts   - Front wheels: IDR 40,000 – 88,000 - Rear wheels: IDR 110,000 – 1,650,000 - Seat: IDR 55,000 – 385,000 - Cushion: IDR 275,000 – 1,100,000 |
| 11 | Shower chairs | None | IDR 880,000 |

Source:

- **Minister of Health Regulation No. 3/2023 on Standard Tariffs for Health Services under the National Health Insurance Programme**
- **Governor of Yogyakarta Special Region Regulation No. 19/2023 on Standard Prices for Health Services under the Universal Health Insurance Programme at the Social Health Insurance Agency of the Yogyakarta Health Office**

Supplementary File 2. UK MRC Process Evaluation Framework for Complex intervention

**Context**

Factors external to the intervention which may influence its implementation, or

whether its mechanisms of impact act as intended.

**Implementation**

Implementation process – the structures, resources and mechanisms through which delivery is achieved

What is delivered

- Fidelity: the consistency of what is implemented with the planned intervention
- Dose: how much intervention is delivered
- Adaptation: changes made in the original
- program during implementation (program modification, reinvention)
- Reach: the extent to which a target audience comes into contact with the intervention

**Mechanisms of impact** – the intermediate mechanisms through which intervention activities produce intended (or unintended) effects

- Participant responses – how participants interact with a complex intervention.
- Mediators – intermediate processes which explain subsequent changes in outcomes.
- Unintended pathways and consequences

Outcomes

Description of intervention and its causal assumption

Source: Moore G, Audrey S, Barker M, *et al.* Process evaluation of complex interventions UK Medical Research Council (MRC) guidance. UK Medical Research Council (MRC). 2015

Supplementary 3. Interview Topic for Each Category of Participants

**Interview Guidelines – Organisation of People with Disabilities (OPDs) in Yogyakarta Province (DIY – abbreviation in local language)**

*Note: Only the primary interview questions are included, and probes have been excluded for conciseness*

Part 1. Initiation and Development of Jamkesus Disabilitas – Policy-making Process

1. Agenda Setting
   a. Why was the issue of disability and access to health insurance raised in DIY?
   b. How was the issue of disability and health insurance for people with disabilities brought up to be made into a regulation/policy in DIY?
   c. Who involved and what were their roles?
   d. What were the challenges and facilitators OPDs faced in raising the issue of disability to get the attention of the public and policymakers?
2. Policy Formulation
   a. How was the process of formulating the policy and regulation on Jamkesus Disability (Governor Regulation No. 51 of 2013 on Special Health Insurance for People with Disabilities) in DIY carried out?
   b. What were the role of OPDs in the policy formulation? Who else involved? What were their roles?
   c. What were the challenges and facilitators in the policy formulation?
3. Policy Adoption
   a. What are the derivate regulations after the initiation of Jamkesus Disabilitas?

Part 2. Implementation of Jamkesus Disabilitas – Policy Implementation

1. What are OPDs role in the implementation of Jamkesus Disabilitas?
2. How is the coordination with the government and implementer?

Part 3. Evaluation and Voices of People with Disabilities

1. What are OPDs role in the monitoring and evaluation of Jamkesus Disabilitas?
2. What are the challenges and facilitators of people with disabilities in accessing Jamkesus Disabilitas?
3. What are the factors that make Jamkesus Disabilitas successful?
4. What need to be improved from Jamkesus Disabilitas?

Part 4. Adoption and Adaptation

1. Why was Jamkesus Disabilitas merged into Jamkesta?
2. If other settings want to adopt Jamkesus Disabilitas, what do they need to prepare?

**Interview Guidelines – People with Disabilities**

*Note: Only the primary interview questions are included, and probes have been excluded for conciseness*

Part 1. Knowledge about Jamkesus Disabilitas

1. What do you know about Jamkesus Disabilitas?
2. How did you know about Jamkesus Disabilitas?

Part 2. Enrolment/registration to Jamkesus Disabilitas

1. What are the procedures to register to Jamkesus Disabilitas?
2. What were the challenges and facilitators to register to Jamkesus Disabilitas?

Part 3. Experience using Jamkesus Disabilitas services

1. Regular services – Primary care & Referral care
   a. What services did you access?
   b. What are the procedures to access the services?
   c. What are the challenges and facilitators accessing the services?
   d. How accessible is the service for you?
2. Assistive technology (AT)
   a. What AT do you need?
   b. How did you get the AT you needed before receiving AT from Jamkesus Disabilitas,?
   c. What were the procedures to get AT within Jamkesus Disabilitas?
   d. How do you usually access repair services within Jamkesus Disabilitas?
   a. What were the challenges and facilitators to access AT service?
2. One stop service
   a. What do you know about “One-stop service”?
   b. Why did you access “one-stop service”?
   c. What were the procedures you underwent when accessing the “one-stop service”?
   d. What were the challenges and facilitators to access “one-stop service”?
   e. How accessible is the “one-stop service” for you?
3. Homecare (only for those who received the service)
   a. Why did you need homecare services?
   b. How did you access homecare services?
   c. Who provided the services?
   d. How often did the care provider come?
   e. How did you find the services? What are the challenges and facilitators?
4. Posbindu Disabilitas
   a. What do you know about Posbindu Disabilitas?
   b. What are the services provided? Who provided care?
   c. What are the benefits?
   d. What are the challenges and facilitators in accessing the service?
   e. How did you find the services? How accessible is it? (infrastructure, information)

Part 5. Perceived impact of Jamkesus Disabilitas

1. What is the impact of Jamkesus Disabilitas that you feel?
2. What do you suggest to improve Jamkesus Disabilitas services?

**Interview Guidelines – Government of Yogyakarta Province (DIY)**

*Note: Only the primary interview questions are included, and probes have been excluded for conciseness*

Part 1. Initiation and Development of Jamkesus Disabilitas – Policymaking Process

1. Agenda Setting
   a. Why did the issue of disability and health become government’s concern?
2. Policy Formulation
   a. What were the process of formulating the policy on disability and on Jamkesus Disability in DIY?
   b. Who were involved and what were their roles?
   c. What were the challenges and facilitators in the policy formulation?
3. Policy adoption and initiation of Jamkesus Disabilitas
   a. How was Jamkesus Disabilitas initiated?
   b. Who were involved and what were their roles?
   c. What were the challenges and facilitators in the initiation?

Part 2. Implementation of Jamkesus Disabilitas – Policy Implementation

1. Jamkesus Disabilitas Implementation
   a. What resources needed to implement Jamkesus Disabilitas? (SOP, organisation, human resources, budget, infrastructures)
   b. How to involve service providers in Jamkesus Disabilitas?
   c. Who are involved in the implementation, and what are their roles?
   d. How to coordinate all actors involved in the implementation?
   e. What are the challenges and facilitators in Jamkesus Disabilitas implementation
2. How to reach people with disabilities as beneficiaries? What are the challenges and facilitators?

Part 3. Evaluation

1. How is Jamkesus Disabilitas monitored and evaluated? How did government use evaluation for programme improvement?
2. What are the impacts of Jamkesus Dsabilitas for health system?
3. What are the factors that make Jamkesus Disabilitas successful?
4. What need to be improved from current Jamkesus Disabilitas?

Part 4. Adoption and adaptation

1. Why was Jamkesus Disabilitas merged into Jamkesta?
2. If other setting wants to adopt Jamkesus Disabilitas, what do they need to prepare?

**Interview Guidelines – Service Providers**

*Note: Only the primary interview questions are included, and probes have been excluded for conciseness*

Part 1. Involvement in Jamkesus Disabilitas

1. How did you initially get involved in Jamkesus Disabilitas as service provider?
2. How did [the authority] prepare you to provide service for people with disabilities?

Part 2. Service delivery

1. What services do you provide? What services did people with disabilities mostly access?
2. How do you provide services for people with disabilities?
3. What are the challenges and barriers providing services for people with disabilities?

Part 3. Claims and reimbursement

1. How is the claim and reimbursement process carried out?
2. What are the challenges and facilitators in claim process?

Part 4. Monitoring and evaluation

1. How do [the authority] monitor and evaluate service delivery you provided?
2. What are the impacts of Jamkesus Disabilitas for you as service providers?
3. What need to improve from Jamkesus Disabilitas?
